# Variation in waiting times by diagnostic category: an observational study of 1951 referrals to a neurology outpatient clinic

**DOI:** 10.1101/2021.01.12.21249644

**Authors:** Fran Biggin, Timothy Howcroft, Quinta Davies, Jo Knight, Hedley Emsley

## Abstract

**Objective:** To investigate the frequency of diagnoses seen among new referrals to neurology outpatient services; to understand how these services are used through exploratory analysis of diagnostic tests and follow-up appointments; and to examine the waiting times between referral and appointment.

**Methods:** Routine data from new NHS appointments at a single consultant-delivered clinic between Sept 2016 and January 2019 were collected. These clinical data were then linked to hospital administrative data. The combined data were assigned diagnostic categories based on working diagnoses to allow further analysis using descriptive statistics.

**Results:** Five diagnostic categories accounted for 62% of all patients seen within the study period, the most common of which was headache disorders. Following a first appointment, 50% of all patients were offered at least one diagnostic test, and 35% were offered a follow-up appointment, with variation in both measures by diagnostic category. Waiting times from referral to appointment also varied by diagnostic category. 65% of patients with a seizure/epilepsy disorder were seen within the 18 week referral to treatment target, compared to 38% of patients with a movement disorder.

**Conclusions:** A small number of diagnostic categories account for a large proportion of new patients. This information could be used in policy decision making to describe a minimum subset of categories for diagnostic coding. We found significant differences in waiting times by diagnostic category, as well as tests ordered, and follow-up offered; further investigation could address causes of variation.

## INTRODUCTION

Neurology services in the UK are overwhelmed and the majority of neurologist time is spent in outpatient clinics. Demand outstrips capacity across the UK, although there is substantial geographical variation. This research has been driven by a need to better understand various aspects of neurology outpatient services, including the frequency of diagnostic categories prompting referral, how services are used, waiting times, and how these aspects vary by diagnostic category. It is perhaps surprising that research is required at all in order to investigate diagnostic categories, but unlike hospital admissions, UK neurology outpatient services have not routinely applied diagnostic coding to outpatient attendances. This undermines attempts to redesign services and optimise access for patients. The absence of outpatient diagnostic coding also prevents research on this theme, including analyses of variation between clinicians and neurological services. Even where diagnostic coding is utilised, the use of different approaches and coding systems limits comparison. We hope that, as well as offering insights into the frequency of diagnostic categories and how diagnostic categories influence investigations, follow up and waiting times, that this research should also provide a foundation from which to start the process of creating a minimum specification for outpatient neurology coding.

As a specialty neurology in the UK has been under much scrutiny over the past 10 years. In 2011 a review by the National Audit Office entitled “Services for people with neurological conditions” highlighted a number of issues within neurological care in the UK.[1] These issues included, but weren’t limited to, varying quality of diagnoses, poorly coordinated care, inequalities in access to care and workforce shortages. Both an update to this review published in 2015 and a parliamentary paper published in 2016 noted that these issues were still ongoing.[2,3] In order to address these issues at a grassroots level, it is necessary to know who is currently visiting neurology services, and how these services are being used.

A 2010 Kings Fund report stated that referral has direct consequences for patients’ experience of care and costs to the health system.[7] Waiting times for referral can be used as an indication of how overburdened a service is. There is currently limited research on referral practices and waiting times for neurology outpatients. However, as referrals to neurology outpatient clinics in the UK often come from patients’ general practitioners (GPs), research based on all-cause referrals (rather than specialty specific research) highlights the difficulties in understanding variation and how best to respond to it. For example, in 1993 Fertig et al. found that although rates of referral varied between practices, this could not be explained by ‘inappropriate referrals’ and concluded that changes to referral guidelines would be unlikely to reduce referral rates.[8] In contrast, a systematic review conducted by Grimshaw et al in 2008 found that 2 strategies were successful in reducing referral rates; guidelines alongside structured referral sheets; and educational interventions by hospital consultants.[9]

Previous research into neurology outpatient visits in the UK has been carried out by Stevens (1988), Hopkins (1989), and Stone (2010).[4–6] These studies all examined the proportions and demographics of patients presenting with different diagnoses, however none of these papers examined waiting times or rates of diagnostic tests and follow-up. Stone et al acknowledged the importance of knowing what onward treatment patients need and we extend our investigation, conducted over a more recent time period, to these areas.[6]

## Specific Objectives of this study

This research uses routine data collected at a neurology outpatient clinic in North West England to:

- describe the proportions of referrals of patients with different diagnostic categories in order to measure relative service use and guide future research and policy;
- analyse the number of diagnostic tests requested and follow-up appointments offered as a measure of ongoing service use;
- examine waiting times for referral in order to identify potential variation in access to services.

## METHODS

### Study Design

This is a retrospective observational study using routinely collected patient data. The proposal underwent ethical review with both the NHS Research Ethics Committee (Ref: 19/NW/0178) and Confidentiality Advisory Group (Ref: 19/CAG/0056) and received approval from the Health Research Authority (HRA) on 30 May 2019 (Ref: 255676). In addition the study underwent ethical review with Lancaster University Faculty of Health and Medicine Research Ethics Committee and obtained approval on 17 June 2019 (Ref: FHMREC18092).

### Setting and Data Collection

We used data from patients referred to, and offered an appointment in, a single consultant-delivered neurology clinic over a period of three and a half years.

Data were recorded at a neurology outpatient clinic at the Royal Preston Hospital (RPH), which is part of the Lancashire Teaching Hospitals NHS Foundation Trust (LTHTR), provider of the Lancashire and South Cumbria regional neurosciences service. The regional service covers a geographically and socio-economically diverse population of approximately 1.6 million residing in urban areas (including the cities and towns of Preston, Chorley, Lancaster, Blackpool, Blackburn and Burnley) and rural areas (the Fylde coast, rural Lancashire and south Cumbria). This clinic is principally a general neurology clinic with some vascular neurology referrals reflecting the subspecialty interest of the consultant, and is dedicated to adult care. No paediatric referrals were included.

Data were collected prospectively from all new appointments held between 18th September 2015 and 9th January 2019. This totalled 2259 appointments of which 1951 were attended and included in this study. These data were then linked to LTHTR’s business intelligence (BI) database. Patients come from three different referral pathways; under the ‘2 week rule’ for suspected CNS cancer;[10] a 2 week urgent referral for first seizure;[11] or on an 18 week referral to treatment (RTT) timeline.[12] Referrals are triaged by consultant neurologists on a rota, and this may lead to variation in prioritisation to urgent appointments.

The data collected during the clinic represent information which is routinely required for consultation, diagnosis and patient management. This includes information on attendance, patient age, gender, principal working diagnosis, diagnostic tests ordered, and whether a follow up appointment was offered.

The data from BI were used to verify the data collected during clinics (gender, age, and attendance) and to add information regarding the source and date of referral.

### Variables

To undertake statistical analysis it was necessary to categorise the principal working diagnosis as the information was recorded in an uncoded free text field. Several systems exist for formally coding diagnoses (for example ICD-10 and SNOMED-CT) however, in the UK these are not routinely used to code neurology outpatient diagnoses. We used the diagnostic categories used by Stone (2010), as they represent the most recent published work on neurology diagnoses and also provide a pragmatic approach. Where more than one diagnosis had been recorded the principal diagnosis was used.

Diagnostic tests were also categorised from a free text field and include requests for central nervous system (CNS) imaging, other imaging, neurophysiology tests and ‘other’ tests (eg lumbar puncture). For the purposes of this analysis CNS imaging included requests for brain, cervical spine, thoracic spine and/or lumbosacral spine.

### Statistical Methods

For analysis we used R Studio (version 1.2.5019).[13] For the analysis of diagnostic categories and patient demographics analysis we used descriptive statistics including means and proportions. Proportions were also used in the analysis of the number of tests and follow up appointments offered. Chi-square tests of independence were used to test the independence of diagnostic tests ordered and follow-up offered from the diagnostic category.

Raincloud plots and smoothed curves of waiting times from referral for each diagnostic category were created for visual comparison.[14] Empirical cumulative distribution functions (ecdf) with kolmogorov-smirnov tests were used to compare selected distributions of waiting times.

### Missing Data

One record was missing the age of the patient and so does not contribute to the calculations of average age. There were 25 appointments with missing information regarding referral dates. These appointments were included in the analysis of diagnostic category, testing and follow-up frequency, but were excluded from the analyses regarding waiting times. We did not consider any special treatment of missing data as the number of missing records is small (around 1%) and no particular pattern of missingness could be detected.

## RESULTS

### Patient Demographics and Diagnostic Category Frequency

During the study period 1951 first appointments were attended. The mean (SD) age of patients overall was 50.0 (18.6) years and varied from 43.2 (18.6) years for seizure/epilepsy to 74.9 (11.7) years for dementia. The overall proportion of females in the study was 0.56 and the proportions ranged from 0.33 (muscle disorder) to 0.77 (multiple sclerosis).

We recorded 17 different diagnostic categories from approximately 1200 unique free text instances, as described in the methods section, and Table 1 shows an overview of these categories. The five most common diagnostic categories accounted for 62% of all diagnoses and comprised headache, seizure/epilepsy, psychological/functional disorders, movement disorders and peripheral nerve/neuromuscular disorders.

**Table 1:** Overview of Study Characteristics. Chi square test of independence of Number of appointments resulting in at least one test and Diagnostic category: p <0.5 x 10 -10. Chi square test of independence of Number of patients offered follow-up appointment and Diagnostic category: p <0.5 x 10 -10. ^*^proportion of column total. ^**^proportion of diagnostic category.

| Diagnostic category | Number of appointments (proportion*) | Mean age (sd) | Proportion female | Number of appointments resulting in at least one test (proportion**) | Number of patients offered follow-up appointment (proportion**) | Average number of tests ordered per appointment |
| --- | --- | --- | --- | --- | --- | --- |
| Headache (all) | 378 (0.19) | 44.4 (16.9) | 0.69 | 165 (0.44) | 41 (0.11) | 0.49 |
| Seizure/Epilepsy | 282 (0.15) | 43.2 (18.6) | 0.41 | 146 (0.52) | 205 (0.73) | 0.82 |
| Psychological/Functional | 189 (0.10) | 44.3 (14.9) | 0.63 | 85 (0.45) | 36 (0.19) | 0.69 |
| Movement Disorders (all) | 180 (0.09) | 61.5 (18.0) | 0.48 | 55 (0.31) | 94 (0.52) | 0.36 |
| Peripheral nerve/neuromuscular | 166 (0.09) | 59.5 (15.3) | 0.49 | 119 (0.72) | 40 (0.24) | 0.84 |
| Spinal Disorders | 98 (0.05) | 60.0 (15.3) | 0.45 | 80 (0.82) | 20 (0.20) | 1.32 |
| Syncope/transient loss of consciousness | 97 (0.05) | 45.8 (18.1) | 0.54 | 58 (0.60) | 19 (0.20) | 1.03 |
| Stroke (all) | 92 (0.05) | 62.1 (16.3) | 0.46 | 42 (0.46) | 38 (0.41) | 0.66 |
| No definite neurological diagnosis | 66 (0.03) | 48.2 (17.5) | 0.58 | 41 (0.62) | 25 (0.38) | 0.97 |
| Multiple Sclerosis/demyelination | 43 (0.02) | 47.4 (14.9) | 0.77 | 21 (0.49) | 34 (0.79) | 1.09 |
| General Medical | 30 (0.02) | 48.3 (18.3) | 0.57 | 14 (0.47) | 4 (0.13) | 0.50 |
| Dementia | 20 (0.01) | 74.9 (11.7) | 0.30 | 9 (0.45) | 9 (0.45) | 0.45 |
| No Diagnosis Made | 12 (0.01) | 49.6 (9.6) | 0.83 | 0 (0.00) | 0 (0.00) | 0.00 |
| Brain Tumour | 10 (0.01) | 69.8 (16.4) | 0.80 | 6 (0.60) | 2 (0.20) | 0.70 |
| Muscle | 9 (0.00) | 47.3 (21.5) | 0.33 | 5 (0.56) | 6 (0.67) | 1.33 |
| Motor Neurone Disease | 8 (0.00) | 67.4 (7.9) | 0.75 | 4 (0.50) | 7 (0.88) | 0.5 |
| Miscellaneous Neurological Disorders | 271 (0.14) | 50.6 (18.8) | 0.62 | 162 (0.60) | 107 (0.39) | 0.83 |

### Tests ordered and follow up offered

Table 1 shows the proportion of appointments that resulted in at least one diagnostic test being ordered; the proportion that resulted in a follow-up appointment being offered; and the average number of tests offered per appointment. Overall 50% of patients were offered at least one test, and 35% of patients were offered a follow-up appointment. In most of the diagnostic categories it was more likely that at least one test was ordered, than a follow-up appointment being offered. This indicates that some patients were offered a test and simultaneously discharged from the outpatient clinic.

Of the five most common diagnostic categories, seizure/epilepsy patients were offered the highest proportion of follow-up appointments (73%). The smallest proportion of follow-up appointments were offered to headache patients (11%), indicating that 89% of patients falling into the diagnostic category of headache disorders were discharged after only one appointment. The proportion of patients for whom a test was requested is more similar; 52% of seizure/epilepsy patients were offered at least one test compared to 44% of headache patients. Of those tests requested, the majority were for CNS imaging: 98% for headache patients and 82% for seizure/epilepsy. This highlights the heterogeneity of patient pathways which depends largely on diagnostic category. Tests of independence were performed to examine the relationship between test request and diagnostic category, and between follow-up appointment and diagnostic category (p-values shown in the legend of table 1). The results confirm that there is a significant association between both of these variables and diagnostic category.

### Waiting time from referral to appointment

Both raw data and smoothed distributions of waiting times for 11 of the 17 diagnostic categories can be seen in Figure 1. The six diagnostic categories with 30 or fewer appointments over the study period, plus individual appointments with waiting times over 40 weeks (n=2) have been excluded from this figure to optimise visualisation. The full results can be seen in supplementary materials (Figure 3). The two vertical dotted lines on Figure 1 show the targets for a 2 week urgent referral pathway for ‘first fit’ and suspected CNS cancer, and the standard 18 week referral to treatment (RTT) target for non-urgent consultant led appointments. This clearly shows that many patients were not seen within the 18 week timeline.

**Figure 1:**
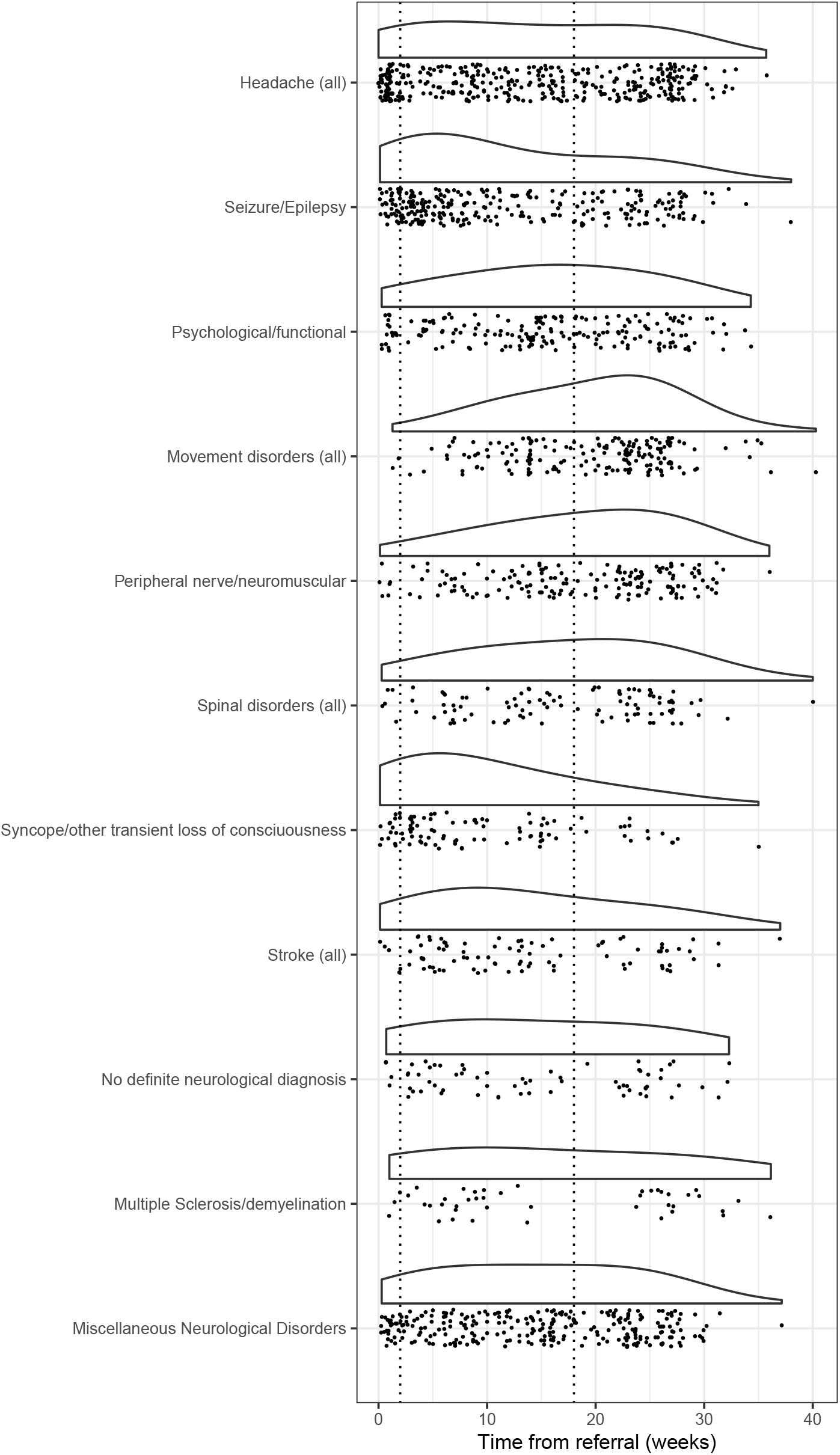
Distribution of waiting time from referral to new appointment.

The proportion of patients referred under two week pathways or the 18 week RTT pathway varied by diagnostic category. Table 2 shows the number and proportion of patients referred on a 2 week urgent pathway for the five most common diagnostic categories, 100% of 2 week headache referrals were on the suspected CNS cancer pathways and 88% of 2 week seizure referrals were on first fit pathway. In order to compare waiting times between diagnostic categories, we identified and removed all referrals made on the 2 week rule or first fit pathway, allowing us to compare routine 18 week referral to treatment only.

**Table 2.** Number and proportion of patients from the 5 most common diagnostic categories referred on a two week pathway (suspected CNS cancer or first seizure), compared to the standard 18 week referral to treatment.

| Diagnostic Category | Number of referrals | Number referred on two week pathways (proportion) | Number referred on standard pathway (proportion) | Unknown (proportion) |
| --- | --- | --- | --- | --- |
| Headache (all) | 378 | 62 (0.16) | 306 (0.81) | 10 (0.03) |
| Seizure/Epilepsy | 282 | 41 (0.15) | 226 (0.80) | 15 (0.05) |
| Psychological/functional | 189 | 13 (0.07) | 170 (0.90) | 6 (0.03) |
| Movement Disorders (all) | 180 | 2 (0.01) | 173 (0.96) | 5 (0.03) |
| Peripheral nerve/neuromuscular | 166 | 5 (0.03) | 158 (0.95) | 3 (0.02) |

Figure 2 shows the empirical cumulative distribution functions (ecdfs) of waiting times for the 5 most common diagnostic categories. The x-axis shows waiting time in weeks and the y-axis represents the proportion of patients who have attended their appointment. Reading along the horizontal dashed line at 0.5 shows the time at which 50% of patients have been seen. Following the vertical line at 18 weeks shows that 65% of seizure/epilepsy patients are seen within target waiting times, compared to only 38% of those diagnosed with a movement disorder.

**Figure 2:**
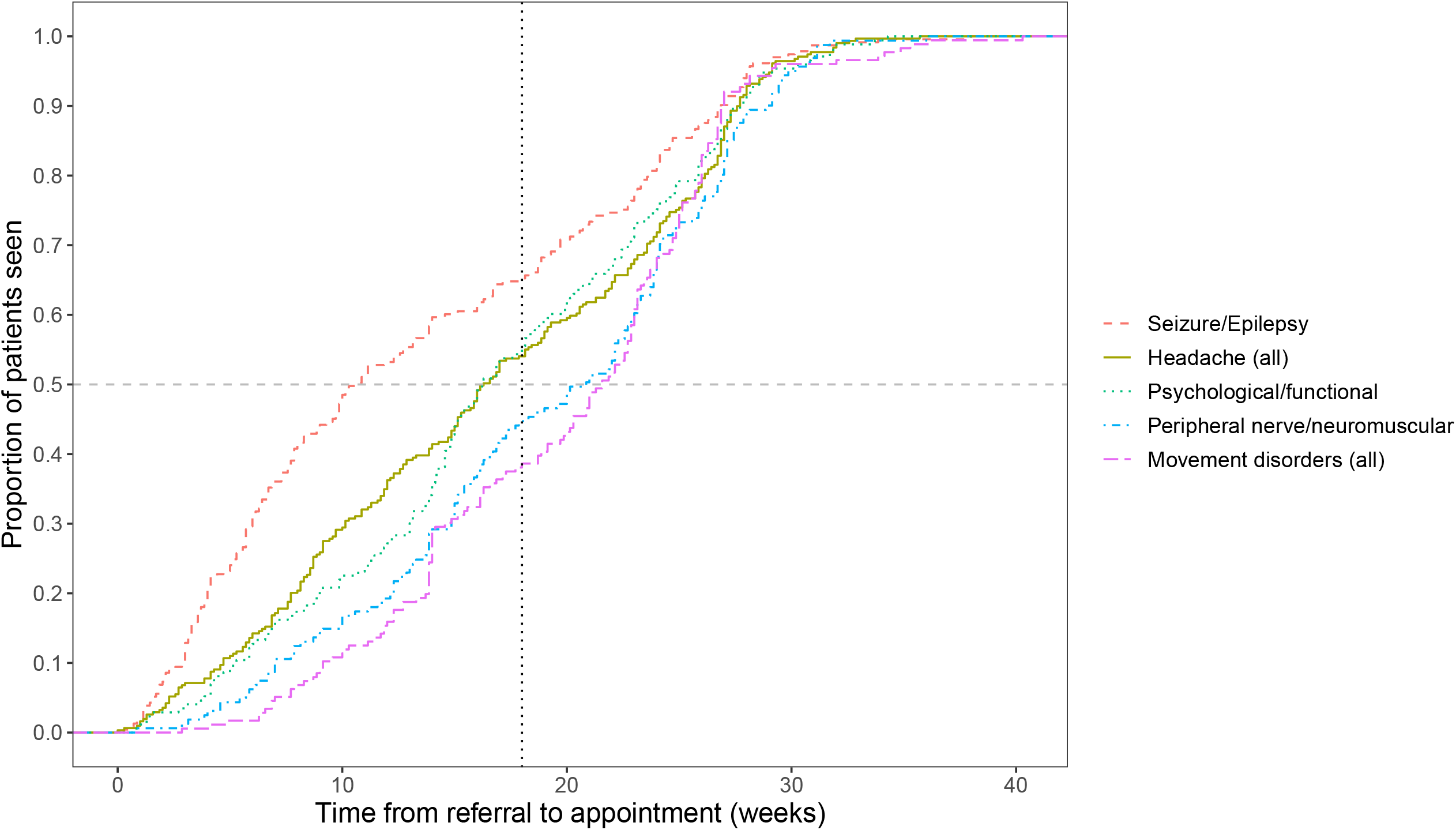
Empirical cumulative distribution of waiting time for the 5 most common diagnostic categories.

Table 3 shows pairwise comparisons of the distributions using the kolmogorov-smirnov test. The size of the p-value indicates the confidence with which we can say the data come from different distributions. Using the Bonferroni correction for multiple testing we need p<0.003 to report a statistically significant difference at *α* = 0.05. Significant p-values are highlighted with an asterisk.

**Table 3.** P-values from pairwise Kolmogorov-Smirnov tests of the empirical cumulative distributions of waiting times for the 5 most common diagnostic categories. * indicates statistically significant values.

|  | Seizure/Epilepsy | Psychological/Functional | Movement Disorders | Peripheral nerve/neuromuscular |
| --- | --- | --- | --- | --- |
| Headache (all) | $2.0 \times 10^{-5} *$ | 0.15 | $1.3 \times 10^{-4} *$ | $4.4 \times 10^{-3}$ |
| Seizure/Epilepsy | | $1.9 \times 10^{-7} *$ | $3.6 \times 10^{-14} *$ | $2.0 \times 10^{-10} *$ |
| Psychological/Functional | | | $3.3 \times 10^{-3}$ | 0.05 |
| Movement Disorders |  |  |  | 0.41 |

## DISCUSSION

This study adds to the current body of research by replicating what previous studies have done in examining frequency of principal working diagnoses and diagnostic categories. We then extended this to look at numbers of diagnostic tests and follow-up appointments offered as a measure of service utilisation, and then compared waiting times from referral to appointment, identifying variations in access to care. Our work is more contemporary by comparison with earlier published work, and has been undertaken during a period impacted by numerous changes in NHS structure and guidance.

### Patient demographics and diagnostic category frequency

The most common diagnostic categories identified in this study were headache, seizure/epilepsy and psychological/functional disorders. This simple but important analysis sheds light on the proportions of patients visiting neurology outpatient clinics falling into headline diagnostic categories. This helps to inform future research and provides valuable information to facilitate service planning and development.

Comparing our results to Stone et al. we see that their four most common diagnostic categories align with four of the five most common diagnostic categories identified by this study; headache, psychological/functional disorders, epilepsy, and peripheral nerve disorders. Despite the fact that the studies were conducted 10 years apart, in different regions of the UK, and using different approaches to data collection - a single consultant in a single centre versus multiple consultants in multiple centres - it is striking that the most common diagnostic categories were similar in both proportion and rank. This points towards a relatively unchanging and predictable list of the most frequent diagnostic categories presenting to UK neurology clinics. This provides an important basis for the coding of outpatient neurology episodes within electronic health records.

### Diagnostic tests ordered and follow up offered

We identify that a large proportion of first appointments result in a diagnostic test being ordered. These tests may be for imaging such as CT or MRI, or neurophysiological tests such as EEG and EMG. They may be requested to provide supportive evidence for a clinical diagnosis or to exclude particular conditions, however many complex factors underpin these requests. Brain imaging requests in particular are surprisingly complex and further discussion is beyond the scope of this work, but it is important to recognise that there are multiple influences beyond direct clinical factors, for example patient reassurance and patient expectations. The need to reassure a patient must be counterbalanced by the potential for incidental findings to provoke anxiety, and it must be acknowledged that patient expectations may be shaped by many influences such as other clinicians, the media, and friends or relatives.

Examining the number of tests ordered and follow-up appointments offered gives a picture of service utilisation which is not shared equally between diagnostic categories. Some diagnostic categories, such as headache, result in a high number of tests, and others such as seizure/epilepsy in a higher proportion of follow-up appointments. This highlights the need for future work into patient pathways in order to examine the way different patients use neurology services.

### Waiting time from referral to appointment

This study shows that waiting times for referral differ by diagnostic category, and that many patients are not seen within the 18 week referral to treatment target. In particular, patients who receive a principal working diagnosis of a movement disorder or a peripheral nerve disorder wait longer on average for their appointment than those with conditions such as headache and seizure/epilepsy. This may be a reflection of the perceived severity and speed of progression of these disorders, and so referrals are made with less urgency. In addition, some variation in prioritisation of referrals as urgent may occur at the point of consultant triage. Referral to a neurology clinic is of often needed in order to assess a patient’s condition, provide a working or definitive diagnosis, and create a plan of care to manage these chronic conditions.[15] Ensuring that the right patients are seen by the right healthcare professional within the most appropriate time frame are key functions of a good referral system,[7] and this study indicates that this may not be happening for those with some disorders.

Waiting times for referral can affect patient satisfaction, interim quality of life, the progression of symptoms, and clinical course.[16] However, more research needs to be conducted into the impact of waiting times on patients referred to neurology services. Although studies have been conducted into ways of streamlining referrals and reducing waiting times, it is currently unknown how or in what way longer waiting times may affect clinical outcomes for patients with neurological conditions.

### Limitations

This study exclusively uses routinely collected data for which there are well established benefits and limitations of its use in research.[17] In the context of this study, the benefit of using routinely collected data lies in its cost-effectiveness, population reach and its reflection of the ‘real world’. Using routinely collected data allows us to see what happens in real time in a clinical population. However, this data is limited in scope, and can suffer from uncertain validity, incompleteness, inaccuracy and inconsistency.[18] For example, because diagnostic coding is not routinely used in neurology outpatient clinics in England the diagnosis information in this study is less reliable than if a standardised system had been used.

In order to overcome these limitations and ensure the data was as accurate as possible, administrative data from the Business Intelligence team was used to verify fields in the data collected from the clinic. This involved linking the data using both NHS and hospital numbers and cross-checking information such as dates of birth, sex and visit dates. Where inconsistencies were found individual records were checked. However, this study is also limited by the unavailability of data that is not collected routinely such as individual socioeconomic status, education level, comorbidities which would help to form a more rounded picture.

The lack of standardised diagnostic coding at neurology outpatient clinics in the UK means that it is exceptionally difficult to carry out a study of this kind across different clinics. In order to enable larger studies a program of coding needs to be established, this would allow data to be collected from different sources. Larger studies would allow greater generalisability as well as comparison between different geographical areas.

Changes in policy and referral practices during the study period may also affect the results, however, we don’t have enough data in this study to determine the possible impact of these changes. Future research could be undertaken to examine key policy changes and their impact on referral times.

### Future Research

This study has opened up many potential avenues for future research. Initially larger studies using data from multiple clinics should be conducted. This would allow for greater generalisability of results and also allow comparison across geographical areas to be made. However, this would be reliant on the introduction of standardised diagnostic coding across the UK.

The identification of the most common diagnostic categories, although unsurprising, may give us the evidence needed to target research to areas which will potentially benefit large groups of patients. This could be directly through innovative approaches to managing common conditions, or indirectly by releasing capacity where possible for other conditions, for instance through the use of alternative headache management pathways.

This study gives insight into how many follow-up appointments and tests are offered. Examining what happens at these follow-up appointments and analysing findings from test results would give us deeper insight into how these resources are bring used, and ultimately whether they are the most appropriate option. This research should be coupled with a health economic approach to examine whether different pathways through referral and diagnosis present different costs and benefits.

Although we identified differences in waiting times for different diagnostic categories, it is unclear how experiencing long waiting times may affect clinical outcomes. More research is needed into how different patient groups experience waiting times, and the potential impact those extended times have on prognosis and treatment.

## CONCLUSION

This study has shown that, by examining routinely collected data, we can analyse service use and variations in access to care. We have highlighted the fact that the principal working diagnoses in more than 60% of patients referred to a neurology outpatient clinic fall into one of only five diagnostic categories, and that the most frequent category is headache disorders. This information is likely to be valuable in the development of approaches to outpatient neurology coding. The analysis of the number of tests and follow-up appointments requested following a first appointment highlights the numerous patient pathways through the service which vary by diagnostic category. Waiting times from referral to appointment show marked variation, indicating variable access between diagnostic categories, with those in the seizure/epilepsy diagnostic category much more likely to be seen within the 18 week timelines than those in movement disorder or peripheral nerve disorder diagnostic categories. This highlights a need to ensure greater consistency of access to outpatient neurology care for patients with suspected neurological disorders. These findings contribute to our understanding of how neurology outpatient services are currently being used and highlight potential areas for future research.

## Data Availability

Due to patient data confidentiality and restrictions imposed by the HRA we are unable to directly share the data for this study. Anyone wishing to access this data must apply to do so through the HRA by completing an IRAS application.

## Declarations

## Acknowledgements

This research was supported by the NIHR Lancashire Clinical Research Facility. The views expressed are those of the authors and not necessarily those of the NHS, the NIHR, or the Department of Health.

## Author Contributions

**Fran Biggin:** Conceptualisation, methodology, software, formal analysis, interpretation, writing – original draft, visualisation **Tim Howcroft:** validation, data curation **Quinta Davies:** validation, data curation **Jo Knight:** Conceptualisation, interpretation, writing – review and editing, supervision, funding acquisition. **Hedley Emsley:** Conceptualisation, interpretation, writing – review and editing, supervision. All authors approved the final manuscript.

## Patient Consent

Not required.

## Ethical Approval

HRA approval granted on 30 May 2019 (Ref: 255676). Lancaster University Faculty of Health and Medicine Research Ethics Committee approval granted on 17 June 2019 (Ref: FHMREC18092).

