## Supplemental Tables for "Variation in waiting times by diagnostic category: an observational study of 1951 referrals to a neurology outpatient clinic"

### 1 Supplementary Materials

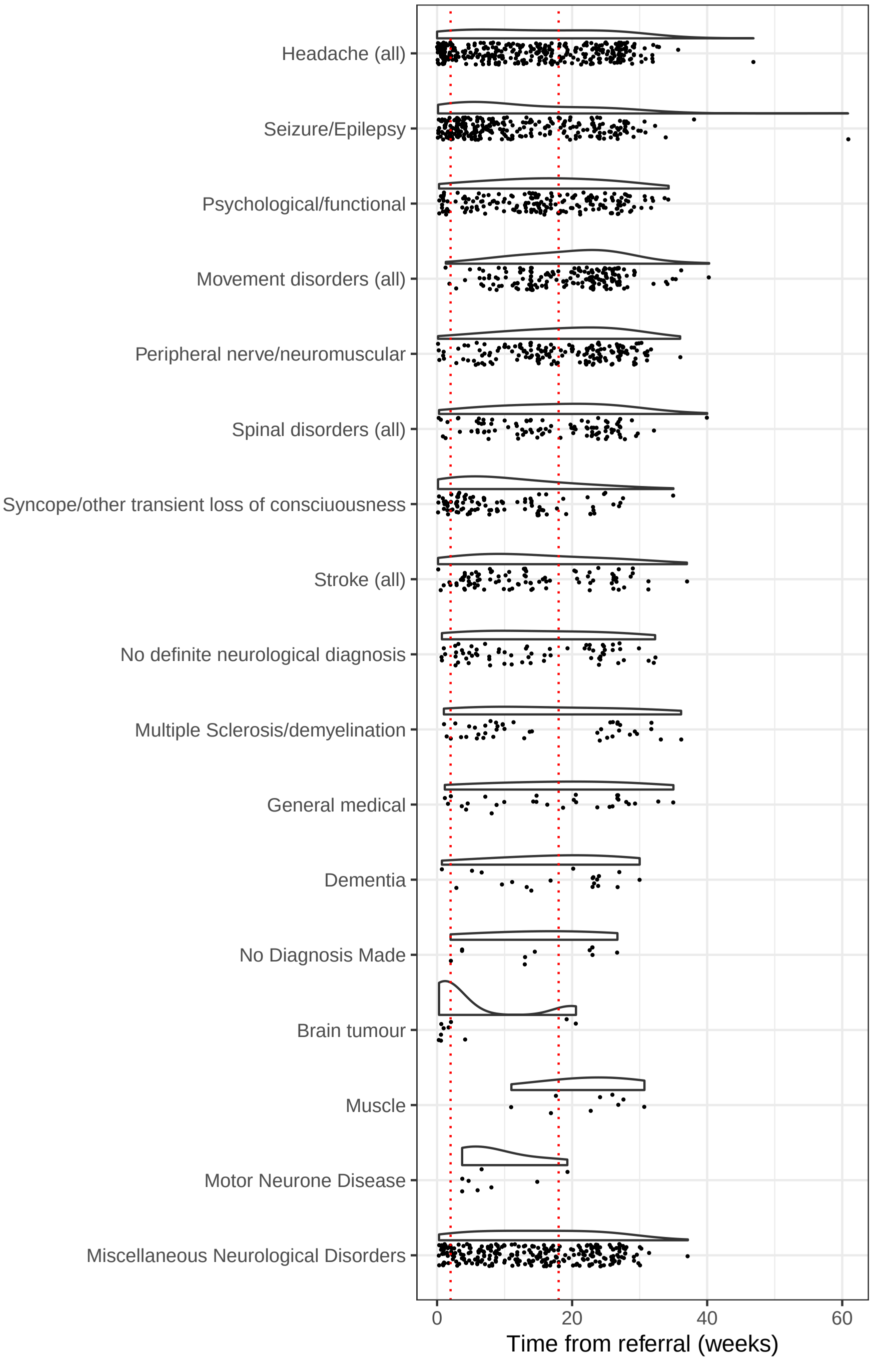


Figure 1: Distribution of waiting time from referral to new appointment including outliers

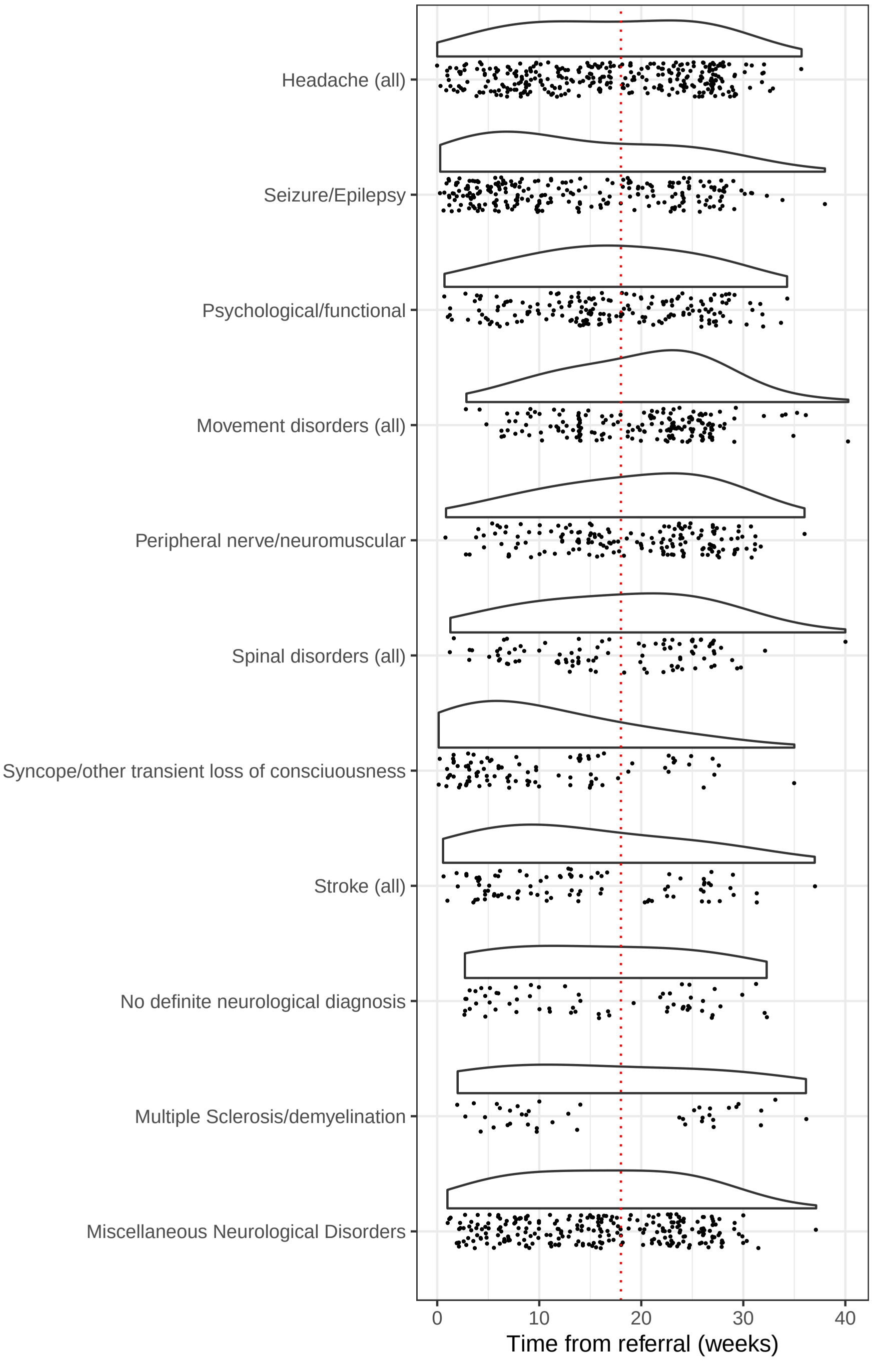


Figure 2: Distribution of waiting time from referral to new appointment after removing patients referred on a 2 week pathway
